# Rapid SARS-CoV-2 antigen detection by immunofluorescence – a new tool to detect infectivity

**DOI:** 10.1101/2020.10.04.20206466

**Authors:** Lorena Porte, Paulette Legarraga, Mirentxu Iruretagoyena, Valeska Vollrath, Gabriel Pizarro, José M. Munita, Rafael Araos, Thomas Weitzel

## Abstract

The evaluated SARS-CoV-2 antigen rapid fluorescence immunoassays reliably identified patients within the first 5 days of symptom onset, when respiratory secretions carried high viral loads. This high performance suggests that these tests might play an important role for future PCR-independent strategies to detect early or infective cases.

## Introduction

Since its emergence in 2019, the SARS-CoV-2 pandemic has resulted in over 30 million confirmed cases and almost 1 million deaths worldwide, as of September 2020 (https://covid19.who.int). Early detection of cases by highly sensitive and specific real-time reverse-transcription polymerase chain reaction (RT-PCR) is the currently recommended diagnostic strategy [1]. However, the high cost of RT-PCR, shortage of reagents, and need for trained personnel have limited the testing capacities of laboratories to provide results in a timely manner [2]. Thus, alternative diagnostic tools allowing the fast testing of large numbers of samples are of high priority [3]. In addition, new aspects of SARS-CoV-2 testing include the ability to evaluate infectivity to help tailor control measures of known or suspected Covid-19 cases [4].

Rapid antigen detection tests (Ag-RDT) using immunochromatographic tests (ICT) or fluorescent immunoassays (FIA) have recently become available; many of which are CE-IVD licensed and some have received FDA emergency use authorization (EUA) [5]. As previously suggested, FIAs are highly specific and can reach remarkably high sensitivities, if applied in samples from early phases of infection or with high viral loads [6,7]. Here we present the performance of two novel FIA automated antigen detection systems in samples from Covid-19 patients presenting within 5 days of symptom onset.

## Material and methods

Samples derived from patients attending Clínica Alemana in Santiago, Chile, for Covid-19 testing. Specimens consisted of naso-oropharyngeal flocked swabs obtained by trained personnel and placed in universal transport media (UTM-RT^®^ System, Copan Diagnostics, Murrieta, CA, USA). Samples were examined for SARS-CoV-2 RNA by RT-PCR assay (COVID-19 Genesig^®^, Primerdesign Ltd., Chander’s Ford, UK). Samples exhibiting exponential amplification curves and cycle thresholds (Ct) values ≤40 were considered positive.

RT-PCR characterized UTM samples were aliquoted and kept at −80° C until analysis by the two FIA kits, “SOFIA SARS Antigen FIA” (Quidel Corporation, San Diego, CA, USA) and “STANDARD^®^ F COVID-19 Ag FIA” (SD Biosensor Inc., Gyeonggi-do, Republic of Korea). Both tests detect SARS-CoV-2 nucleocapsid protein by lateral flow immunofluorescence, which is interpreted by automated analysers (SOFIA 2, Quidel Corporation; F2400, SD Biosensor Inc.). Both kits are CE-IVD labelled; Quidel recently received FDA EUA. Manufacturers state that both tests should be performed using nasopharyngeal swabs collected from symptomatic individuals within 5 days of symptom onset. The use of samples stored in certain brands of transport media (including Copan UTM) is permitted for the SD Biosensor assay; the Quidel test initially also allowed using UTM, but a recent instruction update discourages the use of prediluted samples [8].

For the evaluation, 32 RT-PCR positive UTM samples, all collected within the first 5 days after symptom onset, and 32 negative specimens were selected. All positive samples were from symptomatic patients, 12 negative samples were from asymptomatic patients screened before surgery. Some of the positive (n = 27) and negative samples (n = 19) had been used in a previous evaluation [7]. Assays were performed using the same sample aliquot, following manufacturers’ instructions, by the same laboratory personnel, who were blinded to RT-PCR results. In brief, specimens were mixed with an extraction reagent, dispensed into the cassette’s sample well, and read after incubation by an instrument. All procedures, except the reading, were performed under a BSL2 cabinet. Results were compared to those of RT-PCR as reference method; in case of discordant results, tests were repeated. Demographic and clinical data were obtained from mandatory notification forms and analysed in an anonymized manner. Statistical analysis considered the calculation of sensitivity, specificity, and accuracy using standard formulas, and Wilson score Confidence Interval at 95% (OpenEpi version 3.01). Test performance was evaluated for all samples and for those with high viral loads (Ct ≤25), as previously described [9]. Kits and analysers were provided by manufacturers at reduced costs for evaluation purposes. The study was approved by the institutional review board (Comité Etico Científico, Facultad de Medicina Clínica Alemana, Universidad del Desarrollo, Santiago, Chile) and a waiver of informed consent was granted.

## Results

The study included a total of 64 samples, 32 were RT-PCR positive and 32 RT-PCR negative. The median age was 39 years (IQR 36.7-57) and 52% were male. Median days from symptom onset to RT-PCR testing of positive and negative cases were 2 (IQR 1-3) and 1 (IQR 0.75-4), respectively. Ct values had a median of 17.95 (IQR, 16.4-22.4); 29/32 samples (90.6%) had a Ct ≤25.

Both assays demonstrated an overall sensitivity >90%, reaching 100% for samples with high viral loads (Table 1). False negative results were observed with the Quidel and SD Biosensor assays in two and three samples, respectively, which had Cts of 30.89 to 32.57 and were taken on the fourth or fifth day after symptom onset. Specificity was 96.7% for both tests, i.e. both kits displayed a single false positive result, from two distinct symptomatic RT-PCT negative cases. Both assays were user friendly, included ready-to-use reagents and required little hands-on time. Moreover, analysers were easy-to-use, stored the results, and included options for QR coding, printing, and connection to laboratory information systems.

**Table 1.**
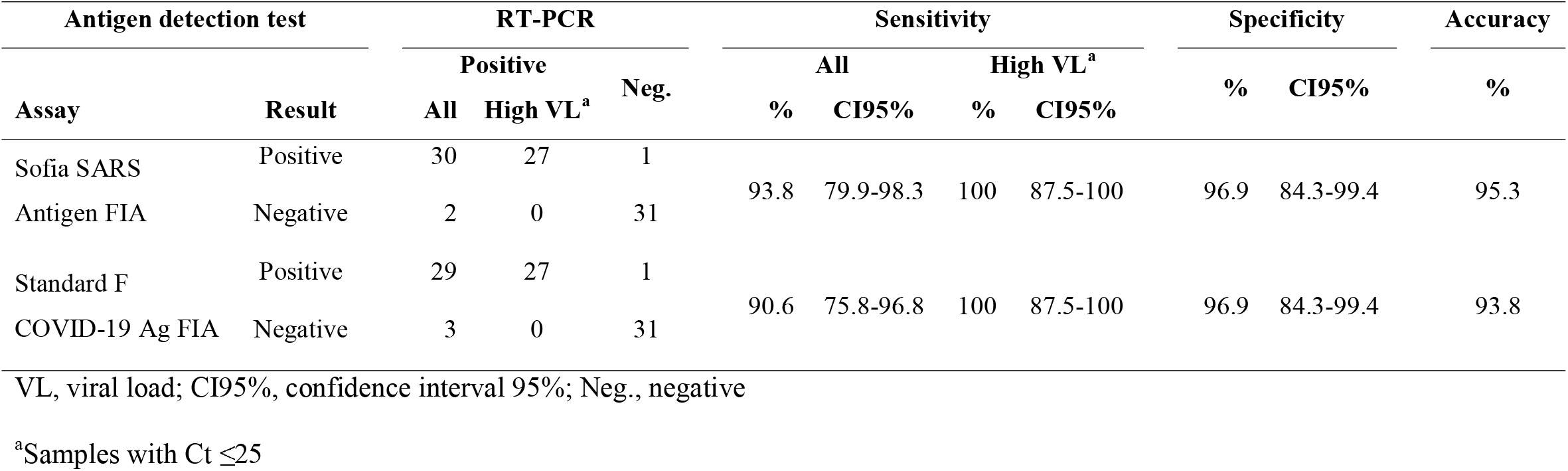
Performance of two automated SARS-CoV-2 antigen detection assays compared to RT-PCR.

## Discussion

At present, RT-PCR is the recommended diagnostic method in patients with suspected SARS-CoV-2 infection [1]. However, material shortages and laboratory capacity limitations, especially during high transmission situations, have caused significant problems and led to the emergence of various new PCR-independent diagnostics [10]. Antigen-based assays are among the most recent developments, but peer-reviewed evaluations of their diagnostic performance are scarce. Hence, their role within the routine diagnostic workup is yet not defined [9,11]. Since antigen detection per se has a lower sensitivity than RT-PCR, it will most likely not replace it [9]. However, the results of this and former studies indicate that antigen detection by immunofluorescence, especially when used with an automated reader, has an excellent sensitivity to detect SARS-CoV-2 in samples with estimated viral loads above ∼10^6^ copies/mL (Ct values ≤25) [9], which are found in pre-symptomatic (1-3 days before symptom onset) and early symptomatic Covid-19 cases (5-7 days after symptom onset) [9,12-14]. According to recent modelling studies, elevated viral titers are associated to infectivity [15]. This is in accordance with *in vitro* experiments, which showed no viral growth from samples with Cts >24 or taken >8 days after symptom onset [16,17]. A viral load of 10^6^ copies/mL has therefore been suggested as the limit of infectivity for clinical practice [18]. However, until the exact threshold of contagiousness is known, other authors have considered a more conservative approach (1,000 copies/mL) [19].

For samples with high viral loads both evaluated tests were 100% sensitive. In our panel of positive samples, false negatives only occurred with Cts >30, which translates to viral loads <10^4^ for the used RT-PCR protocol [20], although this finding has to be confirmed with a larger number of specimens. The high-performance value coincides with recent studies of a similar FIA with automated reading (BioEasy), which demonstrated sensitivities of 100% for samples with Cts ≤25 [6,7] and of 98% for samples with Cts ≤30 [21]. In contrast, immunochromatographic SARS-CoV-2 antigen tests demonstrated lower sensitivity values of 74%-85% for samples with Cts ≤25 [7,22,23].

Although additional studies with larger numbers of samples are needed, the excellent performance data of FIA Ag-RDTs suggest their potential use in the following scenarios, when RT-PCR is unavailable or impractical: 1) closed or semi-closed remote communities such as cruise ships or military camps [9], 2) High-risk congregate facilities including schools, care-homes, dormitories, etc., when testing daily or every other day could reduce secondary infections by 100% or 90%, respectively [24], and 3) screening of asymptomatic attendees at potential superspreader events, like conferences, weddings, and sports or cultural events. In the future, due to their high sensitivity to detect infective patients, FIA Ag-RDTs might also play an important role within “test-out” strategies, i.e. the early release of suspected cases from self-isolation or shortening quarantine for proven cases.

## Data Availability

Data referred to in the manuscript will be available once it is published in a peer-reviewed journal.

## CRediT authorship contribution statement

**L. Porte:** Conceptualization, Data curation, Formal analysis, Investigation, Methodology, Project administration, Supervision, Validation, Writing - original draft, Writing - review & editing. **P. Legarraga:** Formal analysis, Supervision, Validation, Writing - review & editing. **M. Iruretagoyena:** Formal analysis, Validation, Writing - review & editing. **G. Pizarro:** Data curation, Investigation. **V. Vollrath:** Supervision, Validation, Writing - review & editing. **J**.**M. Munita:** Validation, Writing - review & editing. **R. Araos:** Validation, Writing - review & editing. **T. Weitzel:** Conceptualization, Formal analysis, Methodology, Project administration, Validation, Writing - original draft, Writing - review & editing.

